# COMPARING THE VALUE OF DYNAMIC VS. STATIC-IMAGE-BASED TESTS OF EMOTION RECOGNITION IN NEURODEGENERATIVE DISEASES

**DOI:** 10.1101/2024.11.20.24317663

**Authors:** Hulya Ulugut, Tal Shany-Ur, Angelina Quagletti, Faatimah Syed, Bailey McEachen, Joel H. Kramer, Katherine Possin, Bruce L. Miller, Virginia E. Sturm, Maria Luisa Gorno-Tempini, Katherine P. Rankin

## Abstract

**Introduction:** More precise subtyping within dementia syndromes leads to better prediction of pathology, supporting individualized, disease-specific treatments. Notably, studies highlight that identification of the right-temporal or semantic behavioral variant frontotemporal dementia (sbvFTD) subtype relies in part on measuring emotion recognition abilities.

**Methods:** To evaluate the effectiveness of current tools, we compared two dynamic video-based affect labeling tests—the Dynamic Affect Recognition Test (DART) and The Awareness of Social Inference Test-Emotion Evaluation Test (TASIT-EET)—against the static image-based Name Affect subtest of the Comprehensive Affect Testing System (CATS-NA) test. A total of 555 persons with dementia (PwD), in the early stages of neurodegenerative disease (Clinical Dementia Rating ≤ 1; Mini Mental State Examination ≥ 20), diagnosed with Alzheimer’s disease syndrome (AD) (n=154), progressive supranuclear palsy syndrome (PSPS) (n=88), non-fluent variant primary progressive aphasia (nfvPPA) (n=77), semantic variant PPA (n=53), behavioral variant frontotemporal dementia (bvFTD) (n=124), semantic bvFTD (n=65), and 133 healthy older participants underwent emotion testing and structural MRI.

**Results:** All emotion labeling tests differentiated PwD from healthy controls (DART, AUC=0.81; TASIT-EET, AUC=0.84; CATS-NA, AUC=0.72), and FTD with social cognition deficits (sbvFTD, bvFTD, and svPPA) from other PwDs (DART, AUC=0.64; TASIT-EET, AUC=0.66; CATS-NA, AUC=0.63). Dynamic tests outperformed CATS-NA in differentiating sbvFTD from bvFTD and svPPA (DART, AUC=0.79; TASIT-EET, AUC=0.74; CATS-NA, AUC=0.60), whereas DART outperformed TASIT-EET in differentiating sbvFTD from svPPA (DART, AUC=0.73; TASIT-EET, AUC=0.66). Multiple linear regression analysis showed that TASIT-EET performance was predicted by visual memory (Benson-delayed) and verbal semantic (BNT, Animal Fluency) functions (p<0.01) and CATS-NA performance was predicted by visuospatial (CATS-Face matching, Number location) (p<0.001) and executive functions (Modified Trail making speed) (p<0.05), while DART was predicted by only working memory functions (Digit span backward) (p<0.05). DART corresponded to the expected structural anatomy of emotion, including right predominant insula, anterior temporal, and orbitofrontal lobes. While both TASIT-EET and CATS-NA shared that pattern of brain anatomy, TASIT-EET correlated with more left temporal structures than DART, and CATS-NA associated with more dorsal structures than DART. Finally, all emotion labeling tests correlated with real-life empathy deficits measured by a standardized informant-based survey.

**Conclusion:** Tasks showing dynamic audio-visual emotion displays showed better effectiveness for diagnostic differentiation of FTD syndromes than static image-based tasks, and the DART showed better clinical and anatomic precision than the TASIT-EET. Emotion identification deficits are a core feature of dementia syndromes like sbvFTD, but occur in the context of additional cognitive deficits. Therefore, careful selection of tests that reflect the key underlying neural circuits related to emotion, and which minimize demand from other cognitive domains, will result in more accurate diagnoses.

## 1. INTRODUCTION

Frontotemporal dementia (FTD) encompasses a heterogeneous group of neurodegenerative disorders, each characterized by distinct clinical syndromes that predominantly affect behavior and language^1–3^. Notable subtypes of FTD with significant real-life social cognition deficits (SCD) include semantic behavioral variant frontotemporal dementia (sbvFTD) (also known as right temporal FTD), behavioral variant frontotemporal dementia (bvFTD), and semantic variant primary progressive aphasia (svPPA)^1–7^. Among these subtypes, sbvFTD is particularly marked by a deterioration in semantic knowledge related to emotions, resulting in impaired emotion comprehension^5,6^. The quantification of these deficits is essential, not only for accurate diagnosis but also for research investigating the neural mechanisms underlying FTD. Additionally, given the sporadic nature of sbvFTD and its strong association with TDP-43 type C pathology^8–10^, early and precise syndrome diagnosis is crucial for effective patient management, disease monitoring, and appropriate trial registration, as well as improved prediction of disease progression, underlying molecular pathology, and genetic risk factors.

The assessment of socioemotional deficits in clinical practice has traditionally relied on subjective reports from caregivers and clinicians^11^. However, the integration of objective, face-to-face testing during neurological evaluations offers enhanced clarity^12^. Since the pioneering work of Ekman and Friesen^13^, emotion recognition deficits have been widely studied in dementia syndromes using tests in which the participant views single static photographs of a person’s face and is asked to choose the label of the displayed affect from an array of choices^12,14–20^. While these static image-based tests are more helpful than no quantification at all, they possess several limitations. Notably, the stimuli for these tests fail to reflect the dynamic nature of emotional expressions encountered in real life^21,22^. Recent advancements in neuropsychological testing have introduced the use of dynamic video-based assessments, with sound, which offer a more realistic depiction of emotion displays^23–29^. Several groups have demonstrated the superiority of dynamic tests over static tests for more accurately reflecting the underlying neural architecture of emotion reading, calling into question the ecological validity of tests that use static facial expressions and thus lack emotional richness that can be derived from the dynamism of the facial movements, vocal prosody, and body posture components^21,22,30–35^.

Primarily static emotion recognition tests have been extensively utilized in clinical practice and research for FTD, and have demonstrated some effectiveness in distinguishing FTD patients with SCD from healthy controls (HCs) and other persons with dementia (PwD) who do not exhibit social cognition impairments such as Alzheimer’s disease (AD)^6,12,14–18,30,36^. However, it remains unclear the degree to which tests using different types of emotion stimuli are differentially sensitive to discriminate among the FTD subtypes that present with real life socioemotional deficits, and how well test performance corresponds with the brain anatomy known to mediate emotion recognition functions.

The aim of this study is to evaluate the effectiveness of current emotion recognition tools in differentiating among dementia syndromes, and particularly the FTD subtypes with social cognition deficits. By comparing two dynamic video-based tests, the Dynamic Affect Recognition Test (DART) and The Awareness of Social Inference Test-Emotion Evaluation Test (TASIT-EET), and a static image-based test, the Comprehensive Affect Testing System-Name Affect (CATS-NA), we hypothesized that dynamic tests would outperform static tests in accurately differentiating FTD subtypes, and would more precisely correlate with specific brain regions associated with emotion recognition deficits. This study seeks to provide evidence-based recommendations for the selection of neuropsychological tests of emotion reading that best reflect the function of key underlying neural circuits, thereby enhancing diagnostic accuracy and personalized clinical care for individuals with FTD.

## 2. METHODS

### 2.1 Participants

In this study, we evaluated a total of 688 participants, including 555 PwD who had data available from at least one face-to-face emotion recognition test early in their disease course. The sample consisted of 154 individuals with AD syndrome^37^, 88 with progressive supranuclear palsy syndrome (PSPS)^38^, 77 with non-fluent variant primary progressive aphasia (nfvPPA)^3^, 53 with svPPA^3^, 124 with bvFTD^2^, 65 with sbvFTD^4–6^, as well as 133 healthy older participants. All subjects underwent a comprehensive assessment by a multidisciplinary team at University of California San Francisco (UCSF) Memory and Aging Center (MAC). This included structural MRI, neurologic exam, clinical history, and neuropsychological testing in the domains of visuospatial (Benson Figure, CATS Face Matching, Visual Object Space Perception Number Location, MMSE Pentagon), verbal and visual memory (CVLT and Benson Delay), processing speed (Stroop color naming), and executive functioning (Trail making speed, Stroop interference, digit span backwards), as described in our previous studies^39–41^. As part of their recruitment, a caregiver/informant was identified who either lived with the participant or had known them well for more than five years. To obtain the greatest clinical and anatomic precision, we utilized strict inclusion criteria aiming to capture only individuals at the early stages of disease and thus avoiding the more generalized atrophy and globally impaired cognition occurring at later stages of neurodegeneration. Patients were excluded if (i) clinical dementia rating (CDR)^42^ scores were higher than 1, (ii) CDR plus National Alzheimer’s Coordinating Center (NACC) behavior and language domains (FTLD) Sum of Boxes^43^ scores were higher than 8, or (iii) Mini Mental State Examination (MMSE)^44^ scores were lower than 20 (Table 1). Participants in the HC sample were predominantly well-educated English-speaking individuals from the San Francisco Bay Area who had no cognitive or functional deficits and normal neurological exam and MRI scan.

**Table 1:** Demographics and clinical characteristics.

|  | Healthy Control | Persons with Dementia (PwD) |  |  |  |  |  |  |
| --- | --- | --- | --- | --- | --- | --- | --- | --- |
|  |  | OTHER PwD |  |  | SOCIAL COGNITION DISORDERS |  |  |  |
| Diagnostic groups | HC (n=133) | AD (n=154) | nfvPPA (n=77) | PSPS (n=88) | bvFTD (n=124) | svPPA (n=53) | sbvFTD (n=65) | P Value |
| Age | 68.1±8.6 | 66.9±10.7 | 68.7±7.3 | 68.1±8.5 | 62.1±8.2<sup>a,c</sup> | 63.9±8.4 | 67.3±8.4 | <0.001 |
| Sex (M/F) | 60/73 | 80/74 | 30/47 | 43/45 | 84/40<sup>b,d</sup> | 27/26 | 35/30 | 0.02 |
| Education | 17.6±1.4 | 16.6±3.8 | 16.3±1.4 | 16.1±3.2 | 16.4±2.8 | 16.7±2.9 | 16.4±5.6 | 0.28 |
| CDR Global | 0 | 0.7±0.2<sup>a</sup> | 0.3±0.4<sup>a,d</sup> | 0.6±0.3<sup>a</sup> | 0.7±0.3<sup>a</sup> | 0.6±0.3<sup>a</sup> | 0.7±0.3<sup>a</sup> | <0.001 |
| CDR® plus NACC FTLD Global | 0 | 0.9±0.2<sup>a</sup> | 0.7±2.0<sup>a</sup> | 0.91±0.5<sup>a</sup> | 1.4±2.0<sup>a,c</sup> | 0.9±1.5<sup>a</sup> | 1.2±1.5<sup>a</sup> | <0.001 |
| CDR® plus NACC FTLD Sum of Boxes | 0 | 4.8 ±1.7<sup>a</sup> | 3.2±4.3<sup>a</sup> | 4.3±2.7<sup>a</sup> | 6.9±4.2<sup>a,d</sup> | 5.5±3.7<sup>a</sup> | 6.8±3.6<sup>a,c</sup> | <0.001 |
| MMSE | 29.4±0.7 | 24.3±2.3<sup>a,cd</sup> | 26.7±2.5<sup>a</sup> | 26.3±2.9<sup>a</sup> | 26.1±2.9<sup>a</sup> | 25.2±3.0<sup>a</sup> | 25.5±2.9<sup>a</sup> | <0.001 |
| Informant Based Emotion Sensitivity Survey Scores |  |  |  |  |  |  |  |  |
| RSMS | 47.3±8.8 | 36.7±12.9<sup>b</sup> | 39.9±12.3 | 34.5±11.7<sup>b</sup> | 22.9±11.5<sup>a,c,d</sup> | 30.9±14.7<sup>a,d</sup> | 20.6±11.9<sup>a,c,d</sup> | <0.001 |
| Face-to-face Emotion Recognition Test Scores |  |  |  |  |  |  |  |  |
| DART | 10.1±2.1 | 8.7±1.4<sup>b</sup> | 8.2±1.6<sup>b</sup> | 8.1±2.4<sup>b</sup> | 7.8±2.1<sup>a,d</sup> | 7.2±1.7<sup>a,d</sup> | 5.4±2.1<sup>a,c,d</sup> | <0.001 |
| TASIT-EET | 11.6±1.7 | 9.1±2.2<sup>b</sup> | 9.7±1.9<sup>b</sup> | 9.5±2.5<sup>b</sup> | 8.7±2.6<sup>a,d</sup> | 7.6±2.0<sup>a,d</sup> | 6.2±2.3<sup>a,c,d</sup> | <0.001 |
| CATS-NA | 13.2 ±1.5 | 12.4±2.1<sup>b</sup> | 12.1±2.0<sup>b</sup> | 11.1±2.2<sup>b</sup> | 10.6±2.6<sup>a,c</sup> | 11.9±2.1<sup>b</sup> | 10.2±2.7<sup>a,c</sup> | <0.001 |

AD: Alzheimer's disease, nfvPPA: Non-fluent variant primary progressive aphasia, PSPS: Progressive supranuclear palsy syndrome, bvFTD: Behavioral variant frontotemporal dementia, svPPA: Semantic variant primary progressive aphasia, sbvFTD: Semantic behavioral variant frontotemporal dementia, CDR: Clinical dementia rating, CDR plus NACC FTLD: National Alzheimer's Coordinating Center (NACC) behavior and language domains (FTLD), MMSE: Mini mental state examination, RSMS: Revised Self-Monitoring Scale, DART: Dynamic Affect Recognition Test, TASIT-EET: The Awareness of Social Inference Test, Emotion Evaluation Test, CATS-NA: Comprehensive Affect Testing System, Name Affect
a: Different from normal controls at $p < .001$ using Dunnett–Hsu tests.
b: Different from normal controls at $p < .05$ using Dunnett–Hsu tests.
c: Different from other disease group(s) at $p < .001$ using Tukey tests.
d: Different from other disease group(s) at $p < .05$ using Tukey tests.

### 2.2. Behavioral Measures

#### Face to Face Emotion Recognition Tests

##### 2.2.1. a) Comprehensive Affect Testing System, Name Affect (CATS-NA)

The CATS battery consists of 13 subtests which are designed to test facial and vocal affect discrimination, including naming and matching^20^. The CATS Name Affect task (CATS-NA) utilizes the facial emotion stimuli created by Ekman and colleagues, which are comprised of male and female faces posing in the basic emotions generally recognized across cultures, i.e., happiness, sadness, anger, surprise, fear, disgust, and neutral^13,20^. This subtest consists of 16 trials in which the participant is asked to select one option from a set of face-label pairs (i.e., a happy face with the label “happy”, a sad face with the label “sad” etc.) that matches the affective expression of one presented face, and it takes approximately 2 minutes to complete.

##### 2.2.1. b) The Awareness of Social Inference Test, Emotion Evaluation Test (TASIT-EET)

The TASIT-EET is a video-based test measuring participants’ ability to identify the six basic emotions (happy, sad, disgusted, surprised, angry, frightened, and neutral), using videos lasting 15-60 seconds in duration in which actors express emotions through facial, vocal, and gestural modalities^24^. The full version uses 28 videos, 14 (short version) were used for this study. The scripts are neutral in spoken semantic content, i.e. the emotion cannot be determined based on what the actor says, but the actors express the emotion using matched facial, vocal prosody, and gestural cues. After each video, participants are asked to select the correct emotion label from six options^24^. The short version of the TASIT-EET takes approximately 5-7 minutes to complete.

##### 2.2.1. c) The Dynamic Affect Recognition Test (DART)

The DART is a video-based test developed at UCSF^45^. Compared to the TASIT-EET, the DART is shorter (12 items, 20-second video clips), simplifies the scene in each video so that there is always only one actor in front of a simple background with few environmental distractions (many scenes in the EET have two actors on screen), utilizes ethnically diverse actors with American accents (the EET was filmed in Australia and actors have Australian accents). While the EET is only commercially available, the DART is freely available to clinicians and researchers, and can be administered either directly or via the TabCAT cognitive testing platform^46,47^. The DART contains items reflecting the six basic emotions including happy, sad, angry, afraid, surprised, and disgusted, and takes approximately 5 minutes to complete.

#### 2.2.2. Informant Questionnaires

##### 2.2.2 a) Revised Self-Monitoring Scale (RSMS)

The RSMS is a 13-item questionnaire, comprised of two subscales and completed by the participant’s informant. It measures the degree to which individuals are sensitive to the subtle socioemotional behavior expressions of others, and whether they are able to modify their self-presentation on the basis of these social cues. Each question is on a 6 point, Likert scale ranging from “certainly, always false” to “certainly, always true.”^48^.

### 2.3 Brain Behavior Correlation

#### 2.3.1 MRI Acquisition and Voxel Based Morphometry (VBM) Preprocessing

All participants underwent 3-T structural magnetic resonance imaging (MRI) within a maximum of 90 days before or after completing the face-to-face emotion recognition tests using a Magnetom scanner (Siemens Inc., Iselin). All structural T1-weighted images were acquired on 3T-scanners (Siemens Trio and Siemens Prisma) at UCSF. T1-weighted 3D magnetization prepared rapid gradient echo (MPRAGE) sequence was used to obtain the structural images, with acquisition parameters as follows: 160 sagittal slices, 1-mm thick, skip = 0 mm; repetition time = 2300 ms; echo time = 2.98 ms; flip angle = 9°; field of view = 240 × 256mm2; voxel size = 1mm3; matrix size = 256 × 256. The VBM analyses were performed using the open source UCSF Brainsight platform^49^. Structural T1-weighted images were preprocessed using SPM12. The images were visually inspected for artifacts, and underwent bias-correction, segmentation into tissue compartments, and spatial normalization using a single generative model with the standard SPM12 parameters. The default tissue probability maps for grey matter, white matter, cerebrospinal fluid, and all other voxels from SPM12 (TPM.nii) were used^50^. To optimize intersubject registration, each participant’s image was warped to a template derived from 300 confirmed neurologically healthy older adults (ages 44-86, M±SD: 67.2±7.3; 113 males, 186 females) scanned with one of three magnet strengths (1.5T, 3T, 4T), using affine and nonlinear transformations with the help of the diffeomorphic anatomical registration through exponentiated lie algebra (DARTEL) method, with standard implementation in SPM12^51^. In all preprocessing steps, default parameters of the SPM12 toolbox were used. Total volume of each tissue compartment was calculated by applying the modulated, warped and segmented masks for gray matter, white matter, and CSF to the corresponding MWS probability map for that individual, and the total intracranial volume (TIV) was derived by summing the three volumes. The spatially normalized, segmented, and modulated gray matter images were smoothed with an 8-mm FWHM isotropic Gaussian kernel for use in VBM analysis.

### 2.4 Additional Statistical Analyses

Python (v3.5.3) was used to conduct statistical analysis. Diagnostic group differences in age, sex, education, CDR global, CDR plus NACC FTLD global, CDR plus NACC FTLD Sum of the Boxes, MMSE, DART (total score), TASIT-EET (total score), CATS-NA (total score), RSMS (total score) were evaluated using chi-square (for categorical variables) and analysis of variance (ANOVA) (for continuous variables). Tukey post-hoc tests were then conducted to identify differences among diagnostic groups, and Dunnett-Hsu post-hoc tests were used to identify differences between disease groups and controls.

To provide more information about the effectiveness of the CATS-NA, TASIT-EET and DART in making differential diagnostic discriminations, receiver-operating characteristic (ROC) analyses were performed. Area under the curve (AUC) derived from to test the accuracy of each test in PwD vs HC, FTD vs other PwD, sbvFTD vs svPPA, and sbvFTD vs bvFTD.

To determine the predictors of the tests, all of the cognitive test scores listed in section 2.1. were loaded as predictors of a backward stepwise multiple linear regression with either the DART, TASIT-EET or the CATS-NA total score as the outcome variable. To test whether static and dynamic emotion task performances predict observed empathy, we also ran simple linear regression models with informant report of the RSMS as the outcome variable and either the DART, TASIT-EET or CATS-NA as the predictor variable. Gender, age at evaluation were controlled for in all models. Disease severity was controlled by excluding participants with CDR≥2 or MMSE<20, therefore no disease-severity covariates were included in the regressions.

VBM was conducted to identify how the CATS-NA, TASIT-EET and DART total score predicted gray matter volume across all groups in a transdiagnostic model. Linear regressions were used to model the predictive value of the test total score on each voxel’s gray matter volume. In the main analysis, control variables sex, age and total intracranial volume (TIV) were included as covariates of no interest. Family-wise error correction was used to identify the lower bound of the significance threshold for the VBM analysis. In this correction, the maximum t-values of the imaging data compared to behavioral data were re-sampled using the Monte Carlo approach, performing 1000 permutations of the error distribution^52^. The t-value at the 95th percentile of this distribution was taken as the custom cut-off threshold, rendering t-values on or above this cut-off significant at a family-wise error corrected level of p < 0.05^53^.

### Ethical approval

This research was subject to approval by the UCSF Committee for Human Resources Independent Review Board. In all cases, informed consent was gained from the participant or the primary caregiver.

### Data availability

At the time of journal publication, the individual-level data from this study will be shared in a controlled-access repository at the Fair AD Data Initiative Workbench^54^.

## 3. RESULTS

Demographics and clinical characteristics are displayed in Table 1. The bvFTD group was significantly younger than other groups (p<0.001) and had a higher proportion of males (p< 0.05). Dementia severity, as measured by CDR scores, was elevated in all dementia groups compared to HCs (p < 0.001), and bvFTD and sbvFTD showed the highest CDR plus NACC FTLD and NACC FTLD sum of the boxes scores, though these remained in the very mild to mild range. MMSE scores were significantly lower in all dementia groups (p < 0.001), with the AD group scoring the lowest (mean=24.3±standard deviation [SD]=2.3). Emotion sensitivity (RSMS) and emotion recognition (DART, TASIT-EET, CATS-NA) were significantly impaired in PwD groups (p < 0.001), with the bvFTD (DART=7.8±2.1, TASIT-EET=8.7±2.6, CATS-NA=10.6±2.6), svPPA (DART=7.2±1.7, TASIT-EET=7.6±2.0, CATS-NA=11.9±2.1), and particularly sbvFTD (DART=5.4±2.1, TASIT-EET=6.2±2.3, CATS-NA=10.2±2.7) group consistently showing the most pronounced deficits, indicating severe social cognition impairments.

### 3.1. The Effectiveness of the Tests

All emotion recognition tests differentiated PwD from HCs (DART, AUC=0.81; TASIT-EET, AUC=0.84; CATS-NA, AUC=0.72), and the subtypes of FTD known to show social cognition disorders (SCD: sbvFTD, bvFTD, and svPPA) from other PwD (DART, AUC=0.64; TASIT-EET, AUC=0.66; CATS-NA, AUC=0.63). Dynamic tests outperformed CATS-NA in differentiating sbvFTD from bvFTD and svPPA (DART, AUC=0.79; TASIT-EET, AUC=0.74; CATS-NA, AUC=0.60), whereas DART outperformed TASIT-EET in differentiating sbvFTD from svPPA (DART, AUC=0.73; TASIT-EET, AUC=0.66) (Figure 1).

**Figure 1.**
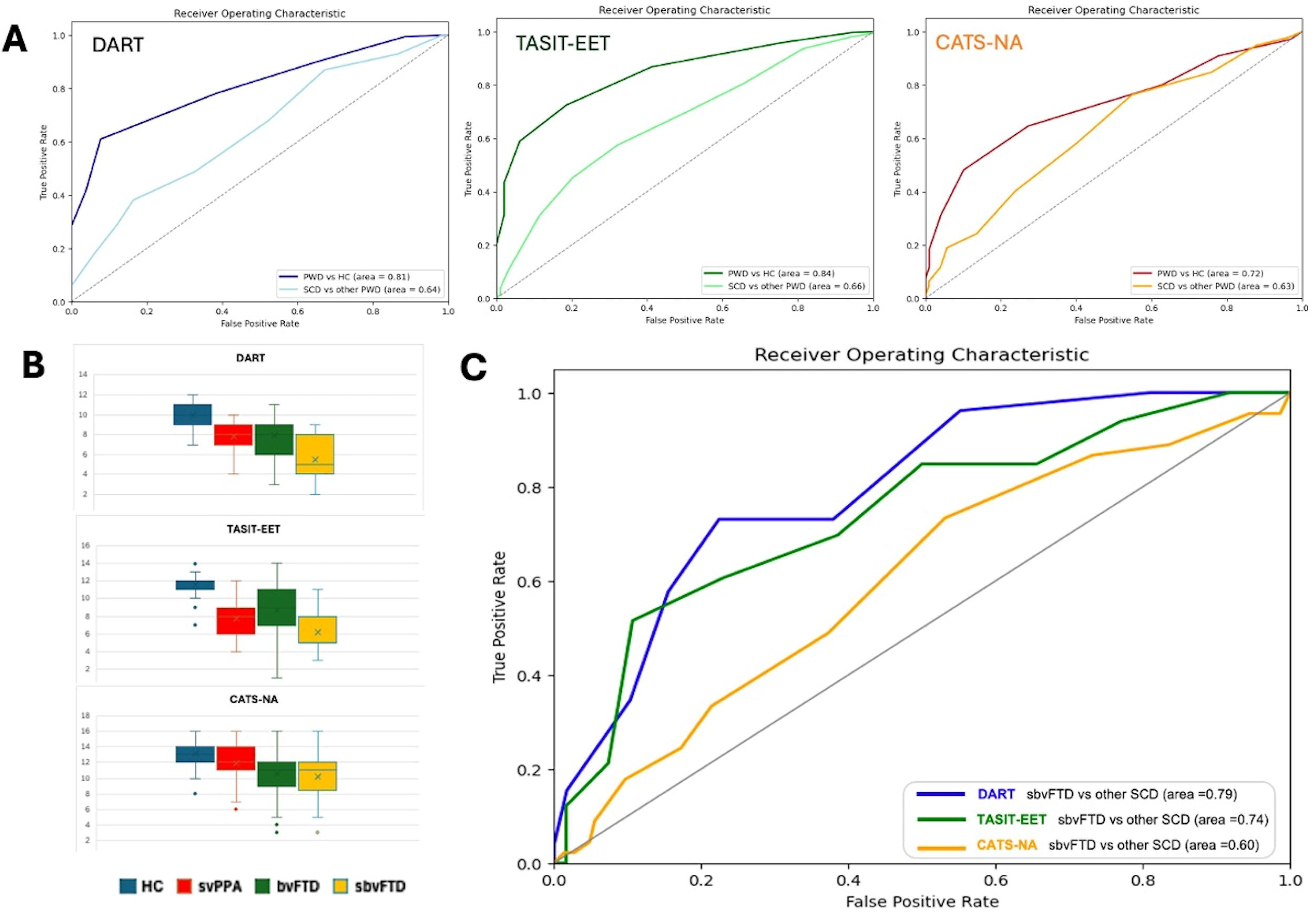
Effectiveness of the emotion recognition tests. **Panel A:** All emotion recognition tests effectively differentiated Persons with Dementia (PwD) from Healthy Controls (HC) and Frontotemporal Dementia (FTD) with Social Cognition Deficits (SCD), including semantic behavioral variant FTD (sbvFTD), behavioral variant FTD (bvFTD), and semantic primary progressive aphasia (svPPA), from other PwD. **Panel B:** Among all FTD with SCD, individuals with sbvFTD exhibited the worst performance across all emotion recognition tests. While patients with bvFTD performed worse on the Comprehensive Affect Testing System-Name Affect (CATS-NA), they demonstrated better performance on The Awareness of Social Inference Test - Emotion Evaluation Test (TASIT-EET). Conversely, patients with svPPA exhibited the opposite pattern, showing worse performance on TASIT-EET but not on CATS-NA. **Panel C:** Dynamic tests, including the Dynamic Affect Recognition Test (DART) and TASIT-EET, outperformed CATS-NA in differentiating sbvFTD from other forms of FTD with SCD.

A subgroup of patients (n=106) underwent all three emotion recognition tests at the same visit. This subgroup analysis demonstrated similar results, showing that sbvFTD exhibited the worst performance among the groups. DART showed the best effectiveness in differentiating sbvFTD from bvFTD and svPPA (AUC=0.73 for both comparisons). TASIT-EET was more effective than CATS-NA for distinguishing sbvFTD from bvFTD (TASIT-EET AUC=0.73, CATS-NA AUC=0.40). However, the difference between the two tests was less pronounced when distinguishing sbvFTD from svPPA (TASIT-EET AUC=0.66, CATS-NA AUC=0.59), while both of them were less effective than the DART for this discrimination (Figure 2).

**Figure 2:**
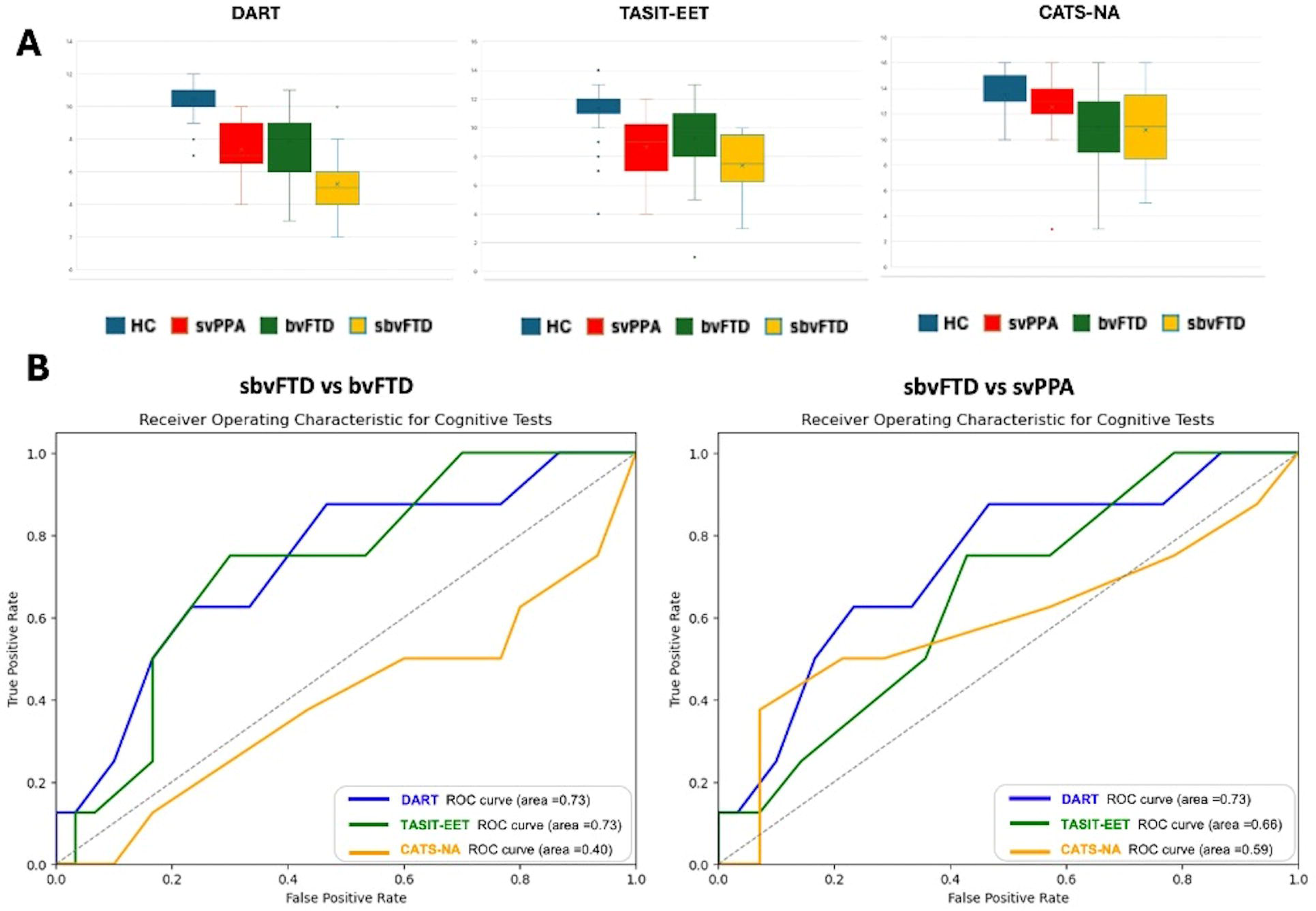
Validation of the results in patients who had all tests at the same visit. **Panel A:** The subgroup analysis focusing on the participants who underwent all three emotion recognition tests at the same visit, demonstrated similar results, showing that semantic behavioral variant frontotemporal dementia (sbvFTD) exhibited the worst performances among other FTD with Social Cognition Deficits (SCD) groups. **Panel B:** The Dynamic Affect Recognition Test (DART) showed the best effectiveness in differentiating sbvFTD from behavioral variant FTD (bvFTD) and semantic variant primary progressive aphasia (svPPA). The Awareness of Social Inference Test - Emotion Evaluation Test (TASIT-EET) showed better performance for distinguishing sbvFTD from bvFTD than distinguishing sbvFTD from svPPA, while the Comprehensive Affect Testing System-Name Affect (CATS-NA) showed the opposite pattern.

### 3.2. Correlation with Empathy Deficits and Predictive Cognitive Functions

All emotion recognition tests were predicted by real-life empathy deficits as reflected by informant-rated RSMS score (p<0.01). Multiple linear regression analyses of neuropsychological test results showed that TASIT-EET performance was significantly predicted by scores on tests of visual memory (Benson-delayed) and verbal semantic (Boston Naming Test, Animal Fluency) functions (p<0.01) while CATS-NA performance was predicted by visuospatial (CATS-Face matching, Number location) (p<0.001) and executive functions (Modified trails Corr/min) (p<0.05). The only significant cognitive predictor for DART performance was the Digit Span Backward test of auditory working memory (p<0.05).

### 3.3. Neuroanatomical Correlates of Emotion Recognition Tests

VBM analysis of emotion recognition tests corrected by age, sex and TIV showed poorer DART performance correlated with focal volume loss in right hemisphere-predominant brain structures including insula, anterior temporal, and orbitofrontal lobes (pFWE<0.05). While both TASIT-EET and CATS-NA shared that same set of core anatomic correlates with the DART, TASIT-EET additionally correlated with left temporal structures including temporal pole, hippocampus and parahippocampus, and CATS-NA associated with more dorsal structures including frontoparietal regions than DART (pFWE<0.05) (Figure 3; Supplementary Table).

**Figure 3:**
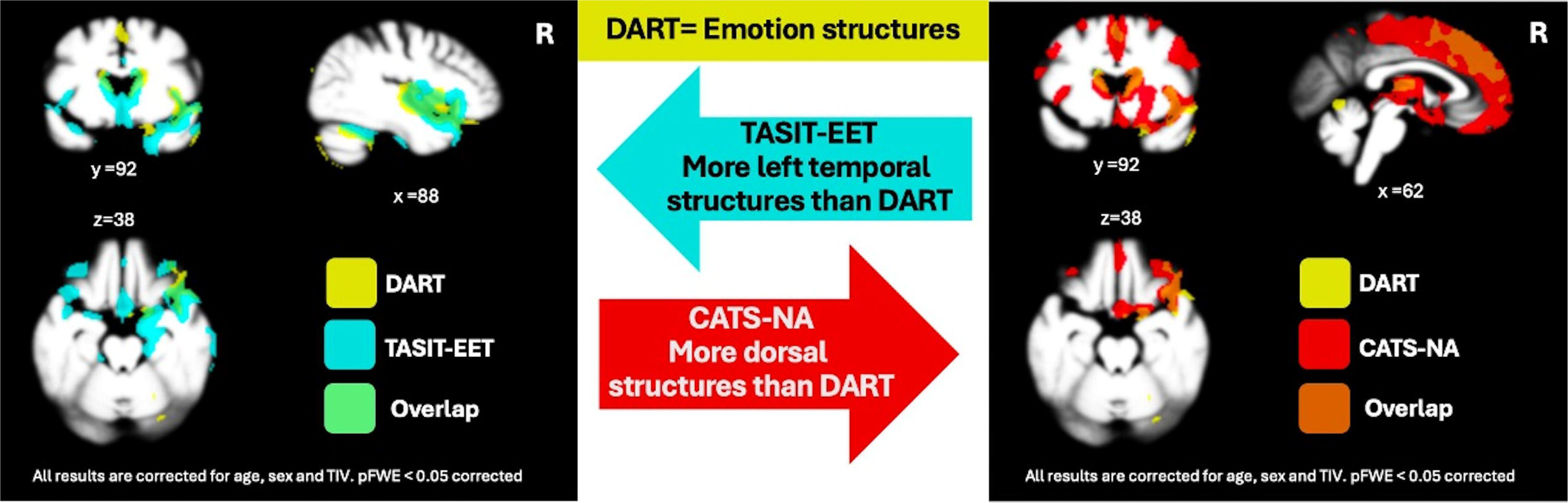
Regional Brain Volume Predicting Test Performance. DART: Dynamic Affect Recognition Test, TASIT-EET: The Awareness of Social Inference Test - Emotion Evaluation Test, CATS-NA: Comprehensive Affect Testing System-Naming Affect

## 4. DISCUSSION

In this study, we evaluated the effectiveness of three emotion labeling tests in PwD, including the static image-based CATS-NA, and the dynamic video-based TASIT-EET and DART. Our findings underscored the importance of dynamic, video-based tests (TASIT-EET, DART) in capturing the emotion recognition deficits which are particularly pronounced in some FTD subtypes, especially in sbvFTD. The DART outperformed TASIT-EET in differentiating sbvFTD from svPPA. More importantly, while the performance on TASIT-EET and CATS-NA was predicted by several cognitive domains outside of emotion reading (visual memory and verbal semantic functions for TASIT-EET, and visuospatial and executive functions for CATS-NA), the DART showed less dependence on functioning in other cognitive domains, being associated only with auditory working memory. Additionally, the DART corresponded focally to the expected structural anatomy mediating emotion reading, including the right predominant insula, anterior temporal, and orbitofrontal lobes. While both TASIT-EET and CATS-NA performance also corresponded with these brain regions, TASIT-EET additionally correlated more with left temporal structures than DART, corresponding to networks mediating semantics, and CATS-NA was associated with more dorsal structures, corresponding to networks mediating executive and visuospatial functioning. Our overall results demonstrated that the DART is a better test for differentiating sbvFTD from other FTD subtypes and from other PwD, and highlighted that the careful selection of tests designed to focally reflect the targeted neural circuits underlying emotion reading, and that minimize demand from other compromised cognitive domains, will lead to more accurate differential diagnosis in PwD.

### 4.1. Clinical Effectiveness of the Tests

Consistent with previous research examining emotion reading in PwD, all three emotion recognition tests—DART, TASIT-EET, and CATS-NA were able to differentiate the FTD subtypes with social cognition deficits from HCs and other PwD effectively.^6,12,14–16,18,30,55–59^ By showing their correspondence with informant-based measures of behavior, our study also confirmed the utility of these tests for reflecting real-world social cognition impairments. However, more recent studies have begun to highlight the centrality of emotion labeling deficits for individuals with sbvFTD^4–6^, even above and beyond the deficits often seen more generally in bvFTD. This is likely because individuals with sbvFTD have focal damage to the right ATL, which is considered to be the hub for semantic knowledge for socioemotional information^60–62^. The international working group for right temporal variant FTD recently highlighted that knowledge loss for emotions is one of the distinct and earliest symptoms of the syndrome which needs to be tested objectively^5^. Our results confirmed the validity of this recommendation by showing that sbvFTD patients exhibited worse performance compared to other PwD on all emotion recognition tests, and that DART exhibits the highest efficacy in distinguishing sbvFTD from bvFTD and svPPA. Although studies have shown that individuals with bvFTD and svPPA also can have emotion recognition deficits, such problems may occur at later stages as disease progresses. Inclusion of only very early-stage patients in our study was an essential study design choice that allowed elucidation of the significant differences in emotion recognition between patients with sbvFTD and those with other FTD with SCD such as bvFTD and svPPA.

Both dynamic tests outperformed static image-based assessments in differentiating sbvFTD from bvFTD in particular. There is some evidence that the underlying mechanisms resulting in poorer performances on the tests differ across syndromes; i.e., they may not be entirely ATL- and semantically-mediated in bvFTD, but involve frontal and subcortical circuitry that mediates emotion experience^6,63–65^. Previous research has shown that the emotion recognition abilities of bvFTD patients improve when emotions are exaggerated and the stimuli are stronger, but decline with subtler expressions or weaker stimuli^66,67^. A neural mechanistic explanation for this result is that the intrinsically connected brain network that is pathognomonically and focally impaired in early bvFTD, i.e. the salience network (SN), plays a role in attention to emotion but not in the evaluation of emotion^63^. This underscores the importance of employing environmentally valid, dynamic stimuli that encompass multiple domains (not only visual facial expressions, but also vocal and body posture cues) to provide a strong, unambiguous signal of emotional expression in the stimuli. A task with these characteristics will better differentiate true emotion identification deficits from errors caused by inattention, or by difficulties created by low fidelity, low signal-to-noise ratio or other ambiguous, attenuated, or conflicting signals in the emotion stimuli themselves.

Although dynamic tests outperformed CATS-NA in differentiating sbvFTD from bvFTD, CATS-NA showed better effectiveness in distinguishing sbvFTD from svPPA, likely due to its language-free nature^20^. DART maintained its effectiveness in differentiating sbvFTD from svPPA as well, as the language component was minimized by removing long conversations and focusing attention by using a single actor in each scene^46^. Our results are consistent with a substantial body of literature noting the significant challenges faced by individuals with svPPA when processing conversations that include longer sentences. Studies have found that in svPPA comprehension declines with increased conversational demands, emphasizing their notable struggles with extended verbal exchanges^68–70^.

### 4.2. Predictive Cognitive Functions

All three emotion recognition tests showed significant association with real-life empathy deficits as measured by a standardized informant-based survey, consistent with the large body of publications showing correspondence between emotion reading test performance and real life behavior^56–58,71,72^. The International Neuropsychological Society (INS) Test Commission emphasizes that an effective test should specifically measure the targeted cognitive function without interference from other cognitive domains, ensuring high specificity^73,74^. This is particularly crucial for PwD, who often experience focal neural dysfunction leading to unique cognitive deficits. Therefore, tests for this population should be brief and practical, avoiding complexities that require prolonged attention spans, intact memory, executive functions, or advanced language skills. In order to examine the potentially confounding influence of additional cognitive deficits on test performance, we identified distinct cognitive predictors for each test. While DART performance was predicted only by working memory functions and not by any other cognitive domain, TASIT-EET performance was significantly associated with visual memory and verbal semantic functions, and CATS-NA performance was linked to visuospatial and executive functions. For the DART and TASIT-EET, participants answer questions on a separate screen after watching short videos, which requires working memory skills^46^. The TASIT-EET specifically features multiple actors in a single scene conducting continuous conversations, and the questions are posed after watching extended videos, thus participants may have additional burden to remember the scene and conversations, even if the script is semantically neutral^24^. The language component in the DART is simplified by minimizing the length, using monologues, and zooming in on actors’ faces and upper bodies without additional distractions. Previous research has demonstrated that understanding dialogue is more challenging compared to monologue, and it depends more significantly on language processing, especially when observing a conversation from a distance, as opposed to focusing on a single person speaking, where attention is directed solely to the individual^68–70,75^. CATS-NA, however, heavily relies on visual skills to decode the facial affect cues and match them with other visual stimuli in the options, ^20^ particularly in the absence of auditory vocal cues, body postures and gestures, and the dynamic movements that provide substantial supportive information while decoding emotions in naturalistic settings. Emotion tests with few extraneous cognitive demands are warranted not only to meet the INS Test Commission criteria but also are particularly suited for intercultural use, because they are more likely to reduce or eliminate reliance on language, incorporate extended and unambiguous displays that are emotionally rich and interculturally clear. Tests like the DART and others that include broader ethnic diversity in the videos are also more ideal for international use^73,74,76^.

### 4.3. Neuroanatomical Correlates

The INS guidelines also indicate that performance on cognitive tests for measuring domain-specific deficits should correlate with function in the expected neural pathways and regions associated with that domain^73^. Such a design ensures the test’s applicability across diverse patient groups with varying cognitive deficits and structural brain changes, and ensures that performance on the test can be interpreted from a neurologic perspective, i.e. as an indicator of focal brain functions. Our neuroimaging analysis revealed that DART corresponds to the expected structural anatomy of emotion, including the predominantly right insula, anterior temporal, and orbitofrontal lobes. The roles of these structures have been demonstrated by many studies, which have shown their association with emotion processing, emotional semantics, and reward-related emotional experiences^60,77–82^. While both TASIT-EET and CATS-NA shared these brain regions, TASIT-EET showed additional correlations with left temporal structures, and CATS-NA was more associated with dorsal brain structures. These results align with previous publications utilizing similar tests, highlighting the involvement of the left temporal structures in language-dominant dynamic emotion recognition tests, and the involvement of the fusiform gyrus, dorsolateral prefrontal cortex, parietal, and occipital lobes in static image-based emotion recognition tests^57,63,83–85^. As discussed in the previous section, the design of the test significantly impacts what brain circuits are recruited to perform the required behavior, as patients may engage different cognitive domains during test performance. These findings underscore the importance of selecting tests that not only measure the targeted cognitive function but also accurately reflect the underlying brain anatomy. Use of tests with more distinct neuroanatomical correlates in research and clinical care is more likely to provide precise and mechanistic insights into emotion recognition deficits in PwD, and particularly those with FTD.

### Limitations and Conclusions

This study provides objective, clinically meaningful evidence about the effectiveness of commonly used tests for differentiating individuals with real-life emotion recognition problems in a large sample of early-stage PwD, by identifying both the cognitive and anatomical correlates of these tests. In our study, dynamic video-based tests, and in particular the DART, were most effective for differential diagnosis across dementia syndromes. Our study includes a large sample size of individuals with sbvFTD, a dementia syndrome for which emotion recognition deficits are pathognomonic, but which is historically excluded in many studies due to the lack of consensus diagnostic criteria. Our sample size is significantly larger than many other studies assessing emotion recognition deficits in this group^6,14,15,18^, enhancing the robustness and reliability of our findings. A relative weakness of our approach was the correlational research design employed; future studies directly examining the functional correlates of test performance by individual PwD in a task-based imaging paradigm would provide important additional information disentangling both cognitive and anatomic correlates of task performance. Emotion identification deficits are a core feature of FTD, particularly in sbvFTD, and careful selection of tests that reflect the key underlying neural circuits, without interference from other cognitive domains, will result in more accurate diagnoses. Future research should focus on validating these findings in more sociodemographically and diagnostically diverse cohorts, as well as exploring the potential of these tools for longitudinal monitoring and intervention development in FTD.

## Data Availability

At the time of journal publication, the individual-level data from this study will be shared in a controlled-access repository at the Fair AD Data Initiative Workbench (https://fair.addi.ad-datainitiative.org/)

## Acknowledgements

Voxel-based morphometry analyses were performed using the UCSF Brainsight system, developed at the UCSF Memory and Aging Center by Katherine P. Rankin, Cosmo Mielke, and Paul Sukhanov, and powered by the VLSM script written by Stephen M. Wilson, with funding from the Rainwater Charitable Foundation and the UCSF Chancellor’s Fund for Precision Medicine. We would furthermore like to thank all participants, their family members and the volunteers that participated in this research, and the project’s research coordinators who helped us with data collection.

## REFERENCES

1. Neary, D. et al. Frontotemporal lobar degeneration: a consensus on clinical diagnostic criteria. Neurology 51, 1546–1554 (1998).

2. Rascovsky, K. et al. Sensitivity of revised diagnostic criteria for the behavioural variant of frontotemporal dementia. Brain 134, 2456–2477 (2011).

3. Gorno-Tempini, M. L. et al. Classification of primary progressive aphasia and its variants. Neurology 76, 1006–1014 (2011).

4. Ulugut, H. et al. Clinical recognition of frontotemporal dementia with right anterior temporal predominance: A multicenter retrospective cohort study. Alzheimers Dement. J. Alzheimers Assoc. 20, 5647–5661 (2024).

5. Ulugut, H. et al. Clinical Recognition of Frontotemporal Dementia with Right Anterior Temporal Predominance; Consensus Recommendations of the International Working Group. medRxiv 2024.10.18.24315786 (2024) doi:10.1101/2024.10.18.24315786.

6. Younes, K. et al. Right temporal degeneration and socioemotional semantics: semantic behavioural variant frontotemporal dementia. Brain 145, 4080–4096 (2022).

7. Rankin, K. P. Brain Networks Supporting Social Cognition in Dementia. Curr. Behav. Neurosci. Rep. 7, 203–211 (2020).

8. Mesulam, M.-M. et al. Frontotemporal Degeneration with Transactive Response DNA-Binding Protein Type C at the Anterior Temporal Lobe. Ann. Neurol. 94, 1–12 (2023).

9. Borghesani, V. et al. Regional and hemispheric susceptibility of the temporal lobe to FTLD-TDP type C pathology. NeuroImage Clin. 28, 102369 (2020).

10. Ulugut, H. et al. Right temporal variant frontotemporal dementia is pathologically heterogeneous: a case-series and a systematic review. Acta Neuropathol. Commun. 9, 131 (2021).

11. Ulugut, H. & Pijnenburg, Y. A. L. Frontotemporal dementia: Past, present, and future. Alzheimers Dement. n/a,.

12. Rankin, K. P. Measuring Behavior and Social Cognition in FTLD. Adv. Exp. Med. Biol. 1281, 51–65 (2021).

13. Ekman, P. & Friesen, W. V. Measuring facial movement. Environ. Psychol. Nonverbal Behav. 1, 56–75 (1976).

14. Kumfor, F. et al. Beyond the face: how context modulates emotion processing in frontotemporal dementia subtypes. Brain 141, 1172–1185 (2018).

15. Kumfor, F. et al. On the right side? A longitudinal study of left-versus right-lateralized semantic dementia. Brain J. Neurol. 139, 986–998 (2016).

16. Bora, E., Velakoulis, D. & Walterfang, M. Meta-Analysis of Facial Emotion Recognition in Behavioral Variant Frontotemporal Dementia: Comparison With Alzheimer Disease and Healthy Controls. J. Geriatr. Psychiatry Neurol. 29, 205–211 (2016).

17. Sturm, V. E. et al. Heightened emotional contagion in mild cognitive impairment and Alzheimer’s disease is associated with temporal lobe degeneration. Proc. Natl. Acad. Sci. U. S. A. 110, 9944–9949 (2013).

18. Bertoux, M. et al. When affect overlaps with concept: emotion recognition in semantic variant of primary progressive aphasia. Brain 143, 3850–3864 (2020).

19. Diehl-Schmid, J. et al. The Ekman 60 Faces Test as a diagnostic instrument in frontotemporal dementia. Arch. Clin. Neuropsychol. Off. J. Natl. Acad. Neuropsychol. 22, 459–464 (2007).

20. Froming, K. B., Ekman, P. & Levy, M. Comprehensive Affect Testing System. 10.1037/t06823-000 (2012).

21. Ferreira, B. L. C., Fabrício, D. de M. & Chagas, M. H. N. Are facial emotion recognition tasks adequate for assessing social cognition in older people? A review of the literature. Arch. Gerontol. Geriatr. 92, 104277 (2021).

22. Roark, D. A., Barrett, S. E., Spence, M. J., Abdi, H. & O’Toole, A. J. Psychological and Neural Perspectives on the Role of Motion in Face Recognition. Behav. Cogn. Neurosci. Rev. 2, 15–46 (2003).

23. Golan, O., Baron-Cohen, S. & Hill, J. The Cambridge Mindreading (CAM) Face-Voice Battery: Testing Complex Emotion Recognition in Adults with and without Asperger Syndrome. J. Autism Dev. Disord. 36, 169–183 (2006).

24. McDonald, S., Flanagan, S., Martin, I. & Saunders, C. The ecological validity of TASIT: A test of social perception. Neuropsychol. Rehabil. 14, 285–302 (2004).

25. Nowicki, S. & Duke, M. P. Individual differences in the nonverbal communication of affect: The Diagnostic Analysis of Nonverbal Accuracy Scale. J. Nonverbal Behav. 18, 9–35 (1994).

26. Scherer, K. R. & Scherer, U. Assessing the ability to recognize facial and vocal expressions of emotion: Construction and validation of the Emotion Recognition Index. J. Nonverbal Behav. 35, 305–326 (2011).

27. Bänziger, T., Mortillaro, M. & Scherer, K. R. Introducing the Geneva Multimodal expression corpus for experimental research on emotion perception. Emotion 12, 1161–1179 (2012).

28. Schlegel, K., Grandjean, D. & Scherer, K. R. Introducing the Geneva Emotion Recognition Test: An example of Rasch-based test development. Psychol. Assess. 26, 666–672 (2014).

29. Mayer, J. D., Salovey, P., Caruso, D. R. & Sitarenios, G. Measuring emotional intelligence with the MSCEIT V2.0. Emotion 3, 97–105 (2003).

30. Goodkind, M. S. et al. Emotion Recognition in Frontotemporal Dementia and Alzheimer’s Disease: A New Film-Based Assessment. Emot. Wash. DC 15, 416– 427 (2015).

31. Rymarczyk, K., Żurawski, Ł., Jankowiak-Siuda, K. & Szatkowska, I. Do dynamic compared to static facial expressions of happiness and anger reveal enhanced facial mimicry? PLoS ONE 11, (2016).

32. Biele, C. & Grabowska, A. Sex differences in perception of emotion intensity in dynamic and static facial expressions. Exp. Brain Res. 171, 1–6 (2006).

33. Trautmann, S. A., Fehr, T. & Herrmann, M. Emotions in motion: dynamic compared to static facial expressions of disgust and happiness reveal more widespread emotion-specific activations. Brain Res. 1284, 100–115 (2009).

34. Sato, W. & Yoshikawa, S. Enhanced experience of emotional arousal in response to dynamic facial expressions. J. Nonverbal Behav. 31, 119–135 (2007).

35. Bucks, R. S. & Radford, S. A. Emotion processing in Alzheimer’s disease. Aging Ment. Health 8, 222–232 (2004).

36. Bertoux, M. et al. Neural correlates of the mini-SEA (Social cognition and Emotional Assessment) in behavioral variant frontotemporal dementia. Brain Imaging Behav. 8, 1–6 (2014).

37. McKhann, G. M. et al. The diagnosis of dementia due to Alzheimer’s disease: recommendations from the National Institute on Aging-Alzheimer’s Association workgroups on diagnostic guidelines for Alzheimer’s disease. Alzheimers Dement. J. Alzheimers Assoc. 7, 263–269 (2011).

38. Höglinger, G. U. et al. Clinical diagnosis of progressive supranuclear palsy: The movement disorder society criteria. Mov. Disord. Off. J. Mov. Disord. Soc. 32, 853– 864 (2017).

39. Gorno-Tempini, M. L. et al. Cognitive and Behavioral Profile in a Case of Right Anterior Temporal Lobe Neurodegeneration. Cortex 40, 631–644 (2004).

40. Possin, K. L., Laluz, V. R., Alcantar, O. Z., Miller, B. L. & Kramer, J. H. Distinct neuroanatomical substrates and cognitive mechanisms of figure copy performance in Alzheimer’s disease and behavioral variant frontotemporal dementia. Neuropsychologia 49, 43–48 (2011).

41. Rijpma, M. G. et al. Semantic knowledge of social interactions is mediated by the hedonic evaluation system in the brain. Cortex J. Devoted Study Nerv. Syst. Behav. 161, 26–37 (2023).

42. Morris, J. C. The Clinical Dementia Rating (CDR): current version and scoring rules. Neurology 43, 2412–2414 (1993).

43. Miyagawa, T. et al. Utility of the global CDR® plus NACC FTLD rating and development of scoring rules: Data from the ARTFL/LEFFTDS Consortium. Alzheimers Dement. 16, 106–117 (2020).

44. Folstein, M. F., Folstein, S. E. & McHugh, P. R. ‘Mini-mental state’. A practical method for grading the cognitive state of patients for the clinician. J. Psychiatr. Res. 12, 189–198 (1975).

45. Rankin, K. P., et al. The Dynamic Affect Recognition Test: Construction and Validation in Neurodegenerative Syndromes. medRxiv. (2024).

46. TabCAT. Memory and Aging Center https://memory.ucsf.edu/research-trials/professional/tabcat.

47. TabCAT Detect cognitive changes earlier. TabCAT Health https://tabcathealth.com.

48. Lennox, R. D. & Wolfe, R. N. Revision of the Self-Monitoring Scale. J. Pers. Soc. Psychol. 46, 1349–1364 (1984).

49. Ucsf, M. Brainsight. Rankin Lab https://rankinlabucsf.wordpress.com/2023/01/12/brainsight/ (2023).

50. Ashburner, J. & Friston, K. J. Unified segmentation. NeuroImage 26, 839–851 (2005).

51. Ashburner, J. & Friston, K. J. Diffeomorphic registration using geodesic shooting and Gauss-Newton optimisation. NeuroImage 55, 954–967 (2011).

52. Dickie, D. A. et al. Permutation and parametric tests for effect sizes in voxel-based morphometry of grey matter volume in brain structural MRI. Magn. Reson. Imaging 33, 1299 (2015).

53. Kimberg, D. Y., Coslett, H. B. & Schwartz, M. F. Power in Voxel-based lesion-symptom mapping. J. Cogn. Neurosci. 19, 1067–1080 (2007).

54. AD Workbench - Login. https://fair.addi.ad-datainitiative.org/.

55. Pressman, P., Gola, K., Shdo, S., Miller, B. & Rankin, K. Relative preservation of affect recognition in posterior cortical atrophy. Neurology 88, (2017).

56. Multani, N. et al. Emotion detection deficits and changes in personality traits linked to loss of white matter integrity in primary progressive aphasia. NeuroImage Clin. 16, 447–454 (2017).

57. Multani, N. et al. Association Between Social Cognition Changes and Resting State Functional Connectivity in Frontotemporal Dementia, Alzheimer’s Disease, Parkinson’s Disease, and Healthy Controls. Front. Neurosci. 13, (2019).

58. Gressie, K. et al. Error profiles of facial emotion recognition in frontotemporal dementia and Alzheimer’s disease. Int. Psychogeriatr. 1–10 (2023) doi:10.1017/S1041610223000297.

59. Park, S. et al. Behavioral and neuroimaging evidence for facial emotion recognition in elderly Korean adults with mild cognitive impairment, Alzheimer’s disease, and frontotemporal dementia. Front. Aging Neurosci. 9, (2017).

60. Binney, R. J. & Ramsey, R. Social Semantics: The role of conceptual knowledge and cognitive control in a neurobiological model of the social brain. Neurosci. Biobehav. Rev. 112, 28–38 (2020).

61. Rice, G. E., Lambon Ralph, M. A. & Hoffman, P. The Roles of Left Versus Right Anterior Temporal Lobes in Conceptual Knowledge: An ALE Meta-analysis of 97 Functional Neuroimaging Studies. Cereb. Cortex 25, 4374–4391 (2015).

62. Rouse, M. A. et al. Social-semantic knowledge in frontotemporal dementia and after anterior temporal lobe resection. 2024.04.11.24305610 Preprint at 10.1101/2024.04.11.24305610 (2024).

63. Yang, W. F. Z. et al. Resting functional connectivity in the semantic appraisal network predicts accuracy of emotion identification. NeuroImage Clin. 31, 102755 (2021).

64. Ulugut Erkoyun, H., et al. A clinical-radiological framework of the right temporal variant of frontotemporal dementia. Brain J. Neurol. 143, 2831–2843 (2020).

65. Kamminga, J. et al. Differentiating between right-lateralised semantic dementia and behavioural-variant frontotemporal dementia: an examination of clinical characteristics and emotion processing. J. Neurol. Neurosurg. Psychiatry 86, 1082– 1088 (2015).

66. Kumfor, F. et al. Are you really angry? The effect of intensity on facial emotion recognition in frontotemporal dementia. Soc. Neurosci. 6, 502–514 (2011).

67. Oliver, L. D., Virani, K., Finger, E. C. & Mitchell, D. G. V. Is the emotion recognition deficit associated with frontotemporal dementia caused by selective inattention to diagnostic facial features? Neuropsychologia 60, 84–92 (2014).

68. Belder, C. R. S. et al. Primary progressive aphasia: six questions in search of an answer. J. Neurol. 271, 1028 (2023).

69. Suárez-González, A. et al. Semantic Variant Primary Progressive Aphasia: Practical Recommendations for Treatment from 20 Years of Behavioural Research. Brain Sci. 11, 1552 (2021).

70. Taylor-Rubin, C. et al. Communication behaviors associated with successful conversation in semantic variant primary progressive aphasia. Int. Psychogeriatr. 29, 1619–1632 (2017).

71. Franklin, H. D. et al. The Revised Self-Monitoring Scale detects early impairment of social cognition in genetic frontotemporal dementia within the GENFI cohort. Alzheimers Res. Ther. 13, 127 (2021).

72. Toller, G. et al. Revised Self-Monitoring Scale: A potential endpoint for frontotemporal dementia clinical trials. Neurology 94, e2384 (2020).

73. Nguyen, C. M. et al. Neuropsychological application of the International Test Commission Guidelines for Translation and Adapting of Tests. J. Int. Neuropsychol. Soc. JINS 30, 621–634 (2024).

74. Fujii, D. E. M. Incorporating Intersectionality in Neuropsychology: Moving the Discipline Forward. Arch. Clin. Neuropsychol. Off. J. Natl. Acad. Neuropsychol. 38, 154–167 (2023).

75. Papageorgiou, S., Stevens, R. & Goodwin, S. The Relative Difficulty of Dialogic and Monologic Input in a Second-Language Listening Comprehension Test. Lang. Assess. Q. 9, 375–397 (2012).

76. Farina, N. et al. Description of the cross-cultural process adopted in the STRiDE (STrengthening Responses to dementia in DEveloping countries) program: A methodological overview. Alzheimers Dement. Amst. Neth. 14, e12293 (2022).

77. Olson, I. R., McCoy, D., Klobusicky, E. & Ross, L. A. Social cognition and the anterior temporal lobes: a review and theoretical framework. Soc. Cogn. Affect. Neurosci. 8, 123–133 (2013).

78. Rolls, E. T. The orbitofrontal cortex and emotion in health and disease, including depression. Neuropsychologia 128, 14–43 (2019).

79. Wager, T. D., Davidson, M. L., Hughes, B. L., Lindquist, M. A. & Ochsner, K. N. Prefrontal-Subcortical Pathways Mediating Successful Emotion Regulation. Neuron 59, 1037–1050 (2008).

80. Skipper, L. M., Ross, L. A. & Olson, I. R. Sensory and semantic category subdivisions within the anterior temporal lobes. Neuropsychologia 49, 3419–3429 (2011).

81. Uddin, L. Q., Nomi, J. S., Hebert-Seropian, B., Ghaziri, J. & Boucher, O. Structure and function of the human insula. J. Clin. Neurophysiol. Off. Publ. Am. Electroencephalogr. Soc. 34, 300–306 (2017).

82. Chang, L. J., Yarkoni, T., Khaw, M. W. & Sanfey, A. G. Decoding the role of the insula in human cognition: functional parcellation and large-scale reverse inference. Cereb. Cortex N. Y. N 1991 23, 739–749 (2013).

83. Yuan, Z. et al. The neural correlation of emotion recognition ability and depressive symptoms–evidence from the HCP database. Front. Psychiatry 13, (2023).

84. Karl, V. & Rohe, T. Structural brain changes in emotion recognition across the adult lifespan. Soc. Cogn. Affect. Neurosci. 18, nsad052 (2023).

85. Adolphs, R., Damasio, H., Tranel, D. & Damasio, A. R. Cortical Systems for the Recognition of Emotion in Facial Expressions. J. Neurosci. 16, 7678 (1996).

