## Supplementary Material for "COMPARING THE VALUE OF DYNAMIC VS. STATIC-IMAGE-BASED TESTS OF EMOTION RECOGNITION IN NEURODEGENERATIVE DISEASES"

**Table 1: Atrophy in structures related to DART performance**

| **maxT** | **ROI** | **x** | **y** | **z** |
| --- | --- | --- | --- | --- |
| 7.47651 | Right PIns posterior insula | 40 | -13 | -4 |
| 7.43286 | Right PP planum polare | 40 | -13 | -6 |
| 7.07546 | Right AIns anterior insula | 42 | 4 | -9 |
| 6.7073 | Right POrG posterior orbital gyrus | 25 | 19 | -22 |
| 6.65314 | Right TMP temporal pole | 40 | 7 | -19 |
| 6.6472 | Right MTG middle temporal gyrus | 66 | -45 | 0 |
| 6.52059 | Right Basal Forebrain | 18 | 3 | -15 |
| 6.46423 | Left PP planum polare | -43 | -10 | -7 |
| 6.45155 | Right MOrG medial orbital gyrus | 18 | 7 | -21 |
| 6.44864 | Left Caudate | -9 | 10 | 13 |
| 6.40972 | Right FuG fusiform gyrus | 36 | -21 | -33 |
| 6.38298 | Right ITG inferior temporal gyrus | 51 | -9 | -39 |
| 6.37771 | Left TMP temporal pole | -48 | 19 | -18 |
| 6.37571 | Left PIns posterior insula | -42 | -10 | -6 |
| 6.37388 | Left Cerebellum Exterior | -28 | -84 | -36 |
| 6.29564 | Left AIns anterior insula | -39 | 6 | -1 |
| 6.19451 | Right STG superior temporal gyrus | 58 | -1 | -13 |
| 6.17181 | Right TTG transverse temporal gyrus | 43 | -15 | 0 |
| 6.16749 | Right Ent entorhinal area | 19 | 3 | -19 |
| 6.07946 | Right Cerebellum Exterior | 27 | -84 | -34 |
| 6.04592 | Left POrG posterior orbital gyrus | -30 | 10 | -21 |
| 6.03004 | Right Caudate | 10 | 16 | 10 |
| 6.00174 | Left FuG fusiform gyrus | -30 | -30 | -27 |
| 5.99743 | Left PHG parahippocampal gyrus | -18 | -16 | -27 |
| 5.98423 | Left STG superior temporal gyrus | -66 | -22 | 6 |
| 5.98072 | Right AnG angular gyrus | 61 | -52 | 16 |
| 5.96529 | Left MOrG medial orbital gyrus | -22 | 18 | -24 |
| 5.88991 | Left Basal Forebrain | -18 | 1 | -15 |
| 5.85494 | Right Amygdala | 16 | -1 | -16 |
| 5.81483 | Left MSFG superior frontal gyrus medial segment | -1 | 43 | 27 |
| 5.7903 | Right MSFG superior frontal gyrus medial segment | 1 | 43 | 28 |
| 5.77534 | Right PT planum temporale | 64 | -15 | 7 |
| 5.77426 | Left Ent entorhinal area | -21 | 0 | -37 |
| 5.75386 | Left ITG inferior temporal gyrus | -39 | -7 | -43 |
| 5.72106 | Left TTG transverse temporal gyrus | -40 | -18 | 4 |
| 5.71604 | Left PT planum temporale | -63 | -21 | 9 |
| 5.70378 | Right LOrG lateral orbital gyrus | 43 | 27 | -16 |
| 5.65112 | Left Thalamus Proper | -3 | -19 | 9 |
| 5.64188 | Right CO central operculum | 40 | 7 | 4 |
| 5.63718 | Right FO frontal operculum | 42 | 9 | 1 |
| 5.58613 | Right SMC supplementary motor cortex | 1 | 24 | 40 |
| 5.57989 | Left MTG middle temporal gyrus | -51 | 1 | -36 |
| 5.52858 | Left SMC supplementary motor cortex | 0 | 22 | 40 |
| 5.50235 | Right Hippocampus | 33 | -36 | -6 |
| 5.48033 | Left Amygdala | -16 | -3 | -16 |
| 5.45781 | Right PHG parahippocampal gyrus | 27 | -12 | -36 |
| 5.44266 | Right Putamen | 19 | 6 | -12 |
| 5.43705 | Right OrIFG orbital part of the inferior frontal gyrus | 46 | 28 | -15 |
| 5.30889 | Right ACgG anterior cingulate gyrus | 1 | 37 | 25 |
| 5.27921 | Right SCA subcallosal area | 10 | 9 | -19 |
| 5.27098 | Right Thalamus Proper | 10 | -6 | 16 |
| 5.2702 | Left CO central operculum | -43 | -9 | 6 |
| 5.22948 | Left FO frontal operculum | -39 | 9 | 3 |
| 5.18202 | Right TrIFG triangular part of the inferior frontal gyrus | 51 | 40 | 3 |
| 5.15691 | Left TrIFG triangular part of the inferior frontal gyrus | -54 | 24 | 1 |
| 5.09258 | Right Accumbens Area | 6 | 9 | -3 |
| 5.08297 | Left ACgG anterior cingulate gyrus | 0 | 37 | 25 |
| 5.07782 | Right MCgG middle cingulate gyrus | 1 | 21 | 24 |
| 5.07256 | Left SFG superior frontal gyrus | -24 | 46 | 34 |
| 4.97895 | Left SCA subcallosal area | -13 | 7 | -21 |
| 4.95391 | Left Putamen | -21 | 3 | -10 |
| 4.89864 | Left MCgG middle cingulate gyrus | 0 | 22 | 22 |
| 4.84699 | Right IOG inferior occipital gyrus | 51 | -75 | 0 |
| 4.83855 | Right MFG middle frontal gyrus | 39 | 25 | 45 |
| 4.79387 | Left OrIFG orbital part of the inferior frontal gyrus | -45 | 21 | -13 |
| 4.755 | Cerebellar Vermal Lobules I-V | 1 | -60 | -4 |

**Table 2: Atrophy in structures related to TASIT-EET performance**

| **maxT** | **ROI** | **x** | **y** | **z** |
| --- | --- | --- | --- | --- |
| 11.0643 | Left PIns posterior insula | -42 | -3 | -12 |
| 10.9138 | Left PP planum polare | -42 | -4 | -12 |
| 10.3353 | Left TMP temporal pole | -33 | 7 | -24 |
| 10.0725 | Left FuG fusiform gyrus | -27 | -10 | -39 |
| 10.0619 | Left POrG posterior orbital gyrus | -30 | 13 | -27 |
| 10.0021 | Left PHG parahippocampal gyrus | -27 | -12 | -37 |
| 9.99425 | Left Ent entorhinal area | -31 | 4 | -22 |
| 9.5074 | Right FuG fusiform gyrus | 30 | -10 | -42 |
| 9.47765 | Left AIns anterior insula | -42 | 4 | -12 |
| 9.30009 | Right PP planum polare | 42 | 4 | -16 |
| 9.29896 | Left Basal Forebrain | -27 | 1 | -16 |
| 9.28407 | Right TMP temporal pole | 42 | 7 | -18 |
| 9.25342 | Left ITG inferior temporal gyrus | -27 | -3 | -42 |
| 9.11361 | Right ITG inferior temporal gyrus | 31 | -6 | -45 |
| 8.88956 | Left Amygdala | -27 | -10 | -15 |
| 8.87898 | Left MTG middle temporal gyrus | -54 | 3 | -31 |
| 8.80818 | Left MOrG medial orbital gyrus | -21 | 6 | -21 |
| 8.7946 | Right Ent entorhinal area | 19 | 1 | -18 |
| 8.70131 | Right PIns posterior insula | 42 | 0 | -12 |
| 8.63158 | Right PHG parahippocampal gyrus | 27 | -12 | -36 |
| 8.58152 | Right Basal Forebrain | 22 | 3 | -16 |
| 8.54142 | Left Hippocampus | -27 | -12 | -16 |
| 8.539 | Right Amygdala | 18 | -1 | -18 |
| 8.32018 | Right SCA subcallosal area | 1 | 10 | -15 |
| 8.18978 | Left STG superior temporal gyrus | -52 | 0 | -18 |
| 8.10338 | Right MTG middle temporal gyrus | 51 | 0 | -37 |
| 7.86743 | Left Putamen | -21 | 3 | -10 |
| 7.83855 | Left Cerebellum Exterior | -42 | -40 | -28 |
| 7.83228 | Left Caudate | -6 | 9 | 0 |
| 7.76385 | Left SCA subcallosal area | -1 | 12 | -7 |
| 7.73847 | Left Accumbens Area | -4 | 9 | -4 |
| 7.72914 | Right Hippocampus | 15 | -6 | -22 |
| 7.35386 | Right AIns anterior insula | 42 | 1 | -6 |
| 7.29824 | Left TTG transverse temporal gyrus | -40 | -18 | 7 |
| 7.23726 | Right POrG posterior orbital gyrus | 31 | 19 | -22 |
| 7.19754 | Left OrIFG orbital part of the inferior frontal gyrus | -39 | 21 | -9 |
| 7.16872 | Left CO central operculum | -42 | -7 | 7 |
| 7.15606 | Right Caudate | 7 | 9 | 0 |
| 7.14166 | Right STG superior temporal gyrus | 54 | -7 | -13 |
| 7.04857 | Right Accumbens Area | 4 | 9 | -9 |
| 6.90537 | Left Pallidum | -25 | -10 | -7 |
| 6.7843 | Left ACgG anterior cingulate gyrus | -3 | 25 | -12 |
| 6.75858 | Left LOrG lateral orbital gyrus | -36 | 34 | -18 |
| 6.72349 | Left GRe gyrus rectus | -10 | 19 | -24 |
| 6.69074 | Left PT planum temporale | -61 | -19 | 10 |
| 6.6847 | Right ACgG anterior cingulate gyrus | -1 | 22 | -12 |
| 6.68185 | Right Putamen | 22 | 3 | -10 |
| 6.61027 | Right MOrG medial orbital gyrus | 18 | 7 | -21 |
| 6.60468 | Left FO frontal operculum | -40 | 9 | 1 |
| 6.602 | Right MFC medial frontal cortex | -1 | 22 | -15 |
| 6.5481 | Left MFC medial frontal cortex | -3 | 21 | -16 |
| 6.45798 | Left Thalamus Proper | -1 | -19 | 12 |
| 6.41531 | Right TTG transverse temporal gyrus | 43 | -15 | 0 |
| 6.397 | Right GRe gyrus rectus | 1 | 22 | -24 |
| 6.36573 | Right Thalamus Proper | 3 | -19 | 12 |
| 6.32393 | Left AOrG anterior orbital gyrus | -31 | 36 | -16 |
| 6.23726 | Left MSFG superior frontal gyrus medial segment | -1 | 36 | 37 |
| 6.04663 | Right CO central operculum | 43 | -6 | 7 |
| 5.9763 | Right MSFG superior frontal gyrus medial segment | 1 | 33 | 31 |
| 5.84291 | Left TrIFG triangular part of the inferior frontal gyrus | -51 | 34 | 9 |
| 5.84168 | Left LiG lingual gyrus | -21 | -40 | -13 |
| 5.767 | Right OrIFG orbital part of the inferior frontal gyrus | 46 | 28 | -15 |
| 5.70153 | Right LOrG lateral orbital gyrus | 43 | 27 | -16 |
| 5.68398 | Right Pallidum | 25 | -9 | -7 |
| 5.62873 | Right Cerebellum Exterior | 42 | -42 | -30 |
| 5.59201 | Left PCgG posterior cingulate gyrus | -12 | -37 | 0 |
| 5.41359 | Right LiG lingual gyrus | 24 | -39 | -13 |
| 5.30888 | Right SMC supplementary motor cortex | 1 | 24 | 42 |
| 5.28214 | Left SMC supplementary motor cortex | -1 | 24 | 42 |
| 5.24103 | Right MCgG middle cingulate gyrus | 1 | -1 | 31 |
| 5.209 | Left MCgG middle cingulate gyrus | 0 | 22 | 22 |
| 5.16983 | Left OpIFG opercular part of the inferior frontal gyrus | -52 | 24 | 21 |
| 5.08263 | Left MFG middle frontal gyrus | -51 | 25 | 21 |
| 5.07748 | Left SFG superior frontal gyrus | -25 | 45 | 34 |
| 4.99802 | Cerebellar Vermal Lobules I-V | -6 | -57 | -4 |
| 4.93382 | Left SMG supramarginal gyrus | -55 | -42 | 43 |
| 4.93359 | Left PO parietal operculum | -36 | -24 | 16 |
| 4.86711 | Right AOrG anterior orbital gyrus | 33 | 39 | -16 |
| 4.86311 | Right FO frontal operculum | 42 | 9 | 1 |
| 4.84978 | Right PCgG posterior cingulate gyrus | 13 | -36 | -1 |
| 4.82733 | Left PoG postcentral gyrus | -58 | -6 | 33 |
| 4.80603 | Left AnG angular gyrus | -40 | -72 | 40 |

**Table 3: Atrophy in structures related to CATS-NA performance**

| **maxT** | **ROI** | **x** | **y** | **z** |
| --- | --- | --- | --- | --- |
| 8.32229 | Right MSFG superior frontal gyrus medial segment | 3 | 25 | 45 |
| 8.24958 | Right SMC supplementary motor cortex | 3 | 24 | 45 |
| 7.93803 | Right OpIFG opercular part of the inferior frontal gyrus | 54 | 25 | 21 |
| 7.68636 | Right TrIFG triangular part of the inferior frontal gyrus | 54 | 27 | 19 |
| 7.68121 | Right MFG middle frontal gyrus | 52 | 25 | 22 |
| 7.66652 | Left ACgG anterior cingulate gyrus | 0 | 34 | 27 |
| 7.61751 | Right ACgG anterior cingulate gyrus | 1 | 28 | 31 |
| 7.61069 | Left SMC supplementary motor cortex | 0 | 24 | 39 |
| 7.56168 | Left MSFG superior frontal gyrus medial segment | 0 | 25 | 39 |
| 7.45157 | Right Caudate | 10 | 7 | 13 |
| 7.24124 | Right Thalamus Proper | 3 | -18 | 7 |
| 7.1218 | Right MCgG middle cingulate gyrus | 1 | 22 | 34 |
| 7.07608 | Left MCgG middle cingulate gyrus | -1 | 12 | 42 |
| 7.0733 | Right OrIFG orbital part of the inferior frontal gyrus | 37 | 25 | -3 |
| 7.02705 | Right AIns anterior insula | 37 | 24 | -3 |
| 6.97523 | Left Thalamus Proper | 0 | -19 | 7 |
| 6.91194 | Right FO frontal operculum | 37 | 25 | 0 |
| 6.62974 | Right POrG posterior orbital gyrus | 36 | 21 | -12 |
| 6.46306 | Right LOrG lateral orbital gyrus | 36 | 24 | -9 |
| 6.38897 | Left SFG superior frontal gyrus | -18 | 27 | 54 |
| 6.38754 | Right Putamen | 22 | 7 | 4 |
| 6.36991 | Right SFG superior frontal gyrus | 27 | 0 | 58 |
| 6.30803 | Left Caudate | -7 | 9 | 10 |
| 6.2604 | Right CO central operculum | 42 | 6 | 3 |
| 6.20543 | Left AIns anterior insula | -39 | 21 | -3 |
| 6.19594 | Right PrG precentral gyrus | 43 | 9 | 31 |
| 6.17044 | Left FO frontal operculum | -39 | 21 | -1 |
| 6.14382 | Right PCgG posterior cingulate gyrus | 3 | -34 | 48 |
| 6.08575 | Left OrIFG orbital part of the inferior frontal gyrus | -37 | 22 | -4 |
| 6.05089 | Right PIns posterior insula | 42 | -6 | 6 |
| 6.02757 | Right AOrG anterior orbital gyrus | 33 | 37 | -16 |
| 6.01476 | Left FRP frontal pole | -24 | 63 | 1 |
| 6.01409 | Left MFG middle frontal gyrus | -25 | 9 | 55 |
| 5.90752 | Right PoG postcentral gyrus | 57 | -4 | 39 |
| 5.85832 | Right MFC medial frontal cortex | 1 | 49 | -12 |
| 5.70619 | Right GRe gyrus rectus | 6 | 39 | -25 |
| 5.67644 | Right SCA subcallosal area | 3 | 9 | -16 |
| 5.6632 | Right Basal Forebrain | 3 | 7 | -15 |
| 5.61005 | Right FRP frontal pole | 25 | 63 | -3 |
| 5.59633 | Right MPrG precentral gyrus medial segment | 4 | -31 | 49 |
| 5.58493 | Left TrIFG triangular part of the inferior frontal gyrus | -52 | 31 | 12 |
| 5.57266 | Right PP planum polare | 45 | -9 | -4 |
| 5.52257 | Left OpIFG opercular part of the inferior frontal gyrus | -55 | 12 | 18 |
| 5.52001 | Right PCu precuneus | 3 | -37 | 48 |
| 5.49156 | Right TMP temporal pole | 45 | 13 | -12 |
| 5.46522 | Right Pallidum | 19 | 4 | 3 |
| 5.46315 | Left PCgG posterior cingulate gyrus | 0 | -33 | 46 |
| 5.46266 | Left MPrG precentral gyrus medial segment | 0 | -27 | 54 |
| 5.39769 | Right MOrG medial orbital gyrus | 9 | 39 | -25 |
| 5.3554 | Left CO central operculum | -39 | 7 | 6 |
| 5.35481 | Left POrG posterior orbital gyrus | -36 | 22 | -9 |
| 5.3295 | Left LOrG lateral orbital gyrus | -34 | 24 | -7 |
| 5.31571 | Left PrG precentral gyrus | -43 | -9 | 52 |
| 5.30041 | Left MFC medial frontal cortex | -1 | 52 | -12 |
| 5.14629 | Right FuG fusiform gyrus | 43 | -49 | -25 |
| 5.13515 | Right SPL superior parietal lobule | 19 | -63 | 60 |
| 5.10776 | Right Cerebellum Exterior | 45 | -52 | -25 |
| 5.02946 | Right TTG transverse temporal gyrus | 45 | -12 | 0 |
| 5.0235 | Right ITG inferior temporal gyrus | 46 | -52 | -25 |
| 5.01679 | Right SMG supramarginal gyrus | 46 | -37 | 51 |
| 4.95301 | Right STG superior temporal gyrus | 64 | -6 | -10 |
| 4.94555 | Right Ent entorhinal area | 18 | 4 | -22 |
| 4.85903 | Right Accumbens Area | 7 | 16 | -3 |
| 4.84325 | Left AOrG anterior orbital gyrus | -31 | 36 | -16 |
| 4.82565 | Left Cerebellum Exterior | -48 | -48 | -39 |
| 4.78148 | Right MTG middle temporal gyrus | 64 | -6 | -12 |
| 4.75864 | Left PIns posterior insula | -40 | -10 | 4 |
| 4.73219 | Cerebellar Vermal Lobules I-V | 7 | -46 | -4 |
| 4.64889 | Left PoG postcentral gyrus | -57 | -7 | 36 |
| 4.62493 | Cerebellar Vermal Lobules VI-VII | 6 | -69 | -12 |
| 4.62022 | Right LiG lingual gyrus | 10 | -45 | -4 |
| 4.60182 | Left PCu precuneus | 0 | -39 | 49 |
| 4.52988 | Right AnG angular gyrus | 39 | -48 | 51 |
